# Combination therapies delay cognitive decline over 10 years in Alzheimer’s NACC participants

**DOI:** 10.1101/2024.01.31.24301055

**Authors:** Yuan Shang, Georgina Torrandell-Haro, Francesca Vitali, Roberta Diaz Brinton

## Abstract

**INTRODUCTION:** Delaying cognitive decline in Alzheimer’s disease can significantly impact both function and quality of life.

**METHODS:** Longitudinal analysis of National Alzheimer’s Coordinating Center (NACC) dataset of 7,653 mild dementia CDR-SB AD participants at baseline with prescriptions for diabetes (DBMD), lipid-lowering (LIPL), anti-hypertensive (AHTN), and non-steroidal anti-inflammatory (NSD) medications over 10 years was evaluated for change in cognitive function relative to non-treated stratified by sex and APOE genotype.

**RESULTS:** Combination therapy of DBMD+LIPL+AHTN+NSD resulted in a 44% / 35% (MMSE/CDR-SB) delay in cognitive decline at 5 years and 47% / 35% (MMSE/CDR-SB) delay at 10 years. Females and APOE4 carriers exhibited greatest cognitive benefit of combination therapy.

**DISCUSSION:** Combination therapies significantly delayed cognitive decline in NACC AD participants at a magnitude comparable to or greater than beta-amyloid immunomodulator interventions. These data support combination precision medicine targeting AD risk factors to alter the course of the disease that persists for a decade.

## 1. Background

Alzheimer’s disease (AD) is a multifactorial and complex brain disease resulting from multiple combinations of risk factors including diabetes [1], hyperlipidemia [2], hypertension [3], and inflammation [4] that are influenced by sex and APOE genotype [5, 6]. Recently FDA-approved immunotherapies targeting amyloid show promise in slowing cognitive decline during the very mild dementia stage of AD [7, 8]. Despite their effectiveness, monoclonal antibodies come with challenges including ARIA / ARIA-E and high costs [9]. While monoclonal antibodies slow the rate of decline, they do not reverse the course of the disease.

AD is preceded by a preclinical phase that can span up to 20 years prior to diagnosis [10]. Multiple risk factors for AD can initiate or contribute to the preclinical stage of AD and ultimately result in AD if not effectively controlled [11]. Based on our Targeted-Risk-AD-Prevention (TRAP) strategy, 364 AD risk factors were identified with diabetes, dyslipidemia, hypertension, and inflammation as principle drivers [12]. These AD risk factors were associated with 629 FDA-approved drugs, 46 of which were identified as high confidence therapies to reduce the risk of AD [12]. Top-ranked therapeutics included glucose-regulators, lipid-lowering, antihypertensives and anti-inflammatory therapeutics [12]. Pathway analysis of the biological signatures of these drug classes revealed that a combinatorial approach to AD could exert greater efficacy compared to a single-target approach [12]. Impact of diabetes, lipid-lowering, antihypertensives and anti-inflammatory drugs for reducing AD risk is well described [13-20], however clinical trials of these therapeutics in non-symptomatic AD patients were not effective in altering the course of the disease [21-34]. However, a subset of responders to these therapeutics was identified within these unsuccessful clinical trials which highlighted the impact of sex and APOE genotype in treatment outcomes [35-37]. Responder analyses of these trials identified APOE4 carriers as the responders of lipid-lowering therapy statins [35] and male APOE4 carriers as responders to diabetes medication [36, 37].

We investigated rate of progression of cognitive decline in real world clinical data within the National Alzheimer’s Coordinating Center (NACC) database to assess the impact of diabetes medications (DBMD), lipid lowering (LIPL), antihypertensive (AHTN), and non-steroidal anti-inflammatory (NSD) therapeutics in participants diagnosed with AD who were using these medications at time of enrollment for their prescribed conditions. Our findings indicated that the use of these drugs, either individually or in combinations, significantly delayed cognitive decline. Moreover, a combination-drug approach targeting multiple systems of biology yielded greater cognitive benefits. Impact of combination therapy on cognitive performance was influenced by non-modifiable risk factors such as sex and APOE genotype [5]. These findings contribute to a precision and personalized strategy for delaying cognitive decline in AD patients, wherein specific combination of FDA-approved drugs with established safety profiles might prove more effective in subpopulations.

## 2. Methods

### 2.1. Data Source

This study was conducted using the National Alzheimer’s Coordinating Center (NACC) database. The NACC database includes longitudinal data from 48,605 participants enrolled in 33 different Alzheimer’s Disease Research Centers (ADRCs) across the United States. This analysis used the Uniform Data Set (UDS) within the NACC database, which contains subject demographics, diagnosis, prescription records and neurological examination findings from 2005 to present (v62, last access September 2023).

### 2.2. Study Design and Variables

Participants were required to have a minimum of 2 visits or more in one of the ADRCs. The first visit was used as index date for the study, i.e., baseline. Diagnosis of AD was determined using the D1 form which was assessed entirely from clinical data (**Table S1**). Subject demographics including age at baseline, sex, race and ethnicity were assessed using the A1 form (**Table S1**). Medication use was self-reported by participants in the dataset, and therapeutics in the categories following categories were included in the study: diabetes medication (DBMD), lipid-lowering drugs (LIPL), non-steroidal anti-inflammatory therapy (NSD) and anti-hypertensive medication (AHTN) (**Table S1, Table S2**). Participants were categorized into specific therapeutic groups based on whether they received treatment with a single drug or a combination of any considered drugs. Each participant was exclusively assigned to one therapeutic group. The control group or non-treated group (none) was defined as participants who were not using any of the selected therapeutics (**Table S2**).

### 2.3. Primary Outcome

The primary outcome for this study was the change in the score on the Mini-Mental State Examination (MMSE) and Clinical Dementia Rating (CDR) Sum of Boxes (CDR-SB) at 5- and 10-year marks compared to baseline. MMSE was selected as the primary outcome of interest due to its widespread use as a screening tool to assess cognitive impairment, commonly employed as part of the comprehensive evaluation process in the diagnosis of AD. MMSE consists of a series of questions and tasks that evaluate mental abilities including orientation, registration, attention and calculation, recall, language, and visual-spatial skills. This test is scored out of 30 points, with a higher score indicating better cognitive function [38]. MMSE results were examined alongside assessments of CDR-SB scores. The use of both CDR and MMSE contributed to more robust findings, as CDR offers a standardized framework to evaluate dementia severity of dementia and categorize individuals across different stages, ranging from normal cognition to severe dementia [39]. This test assesses six cognitive and functional domains including memory, orientation, judgment, home and hobbies, community affairs and personal care. Each domain is scored from 0 to 3 with higher scores indicating worse performance. The CDR-SB score is the cumulative score of each domain and ranges from 0 to 18 with higher scores indicating severe dementia [39]. MMSE and CDR-SB results were extracted from the forms C1 and B4 of the NACC database, respectively (**Table S1**). Drug groups with insufficient participant number in the sex groups (n<50) were excluded from these analysis (DBMD alone, DBMD+AHTN, DBMD+LIPL, DBMD+NSD, **Table 1**).

**Table 1:**
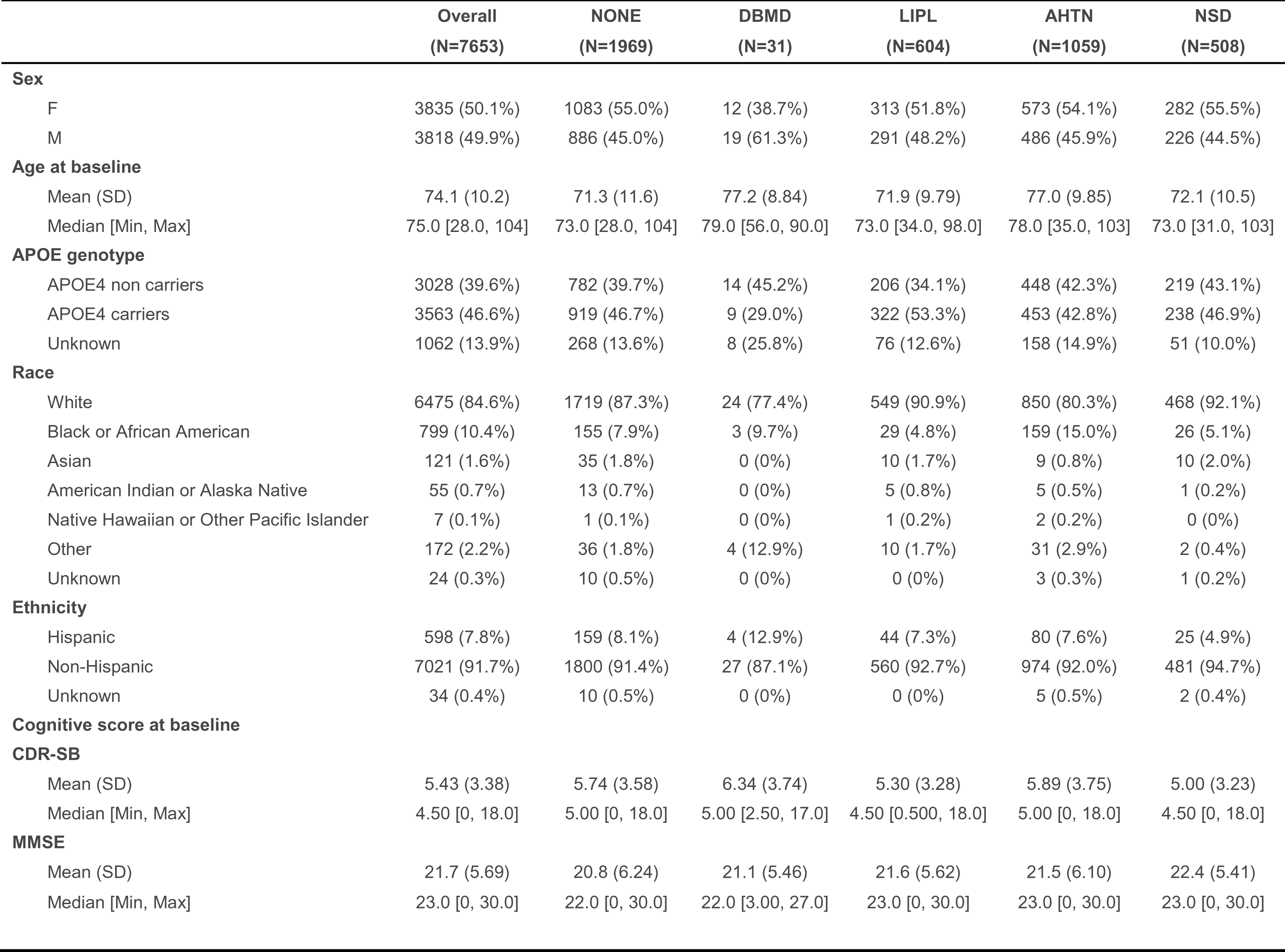
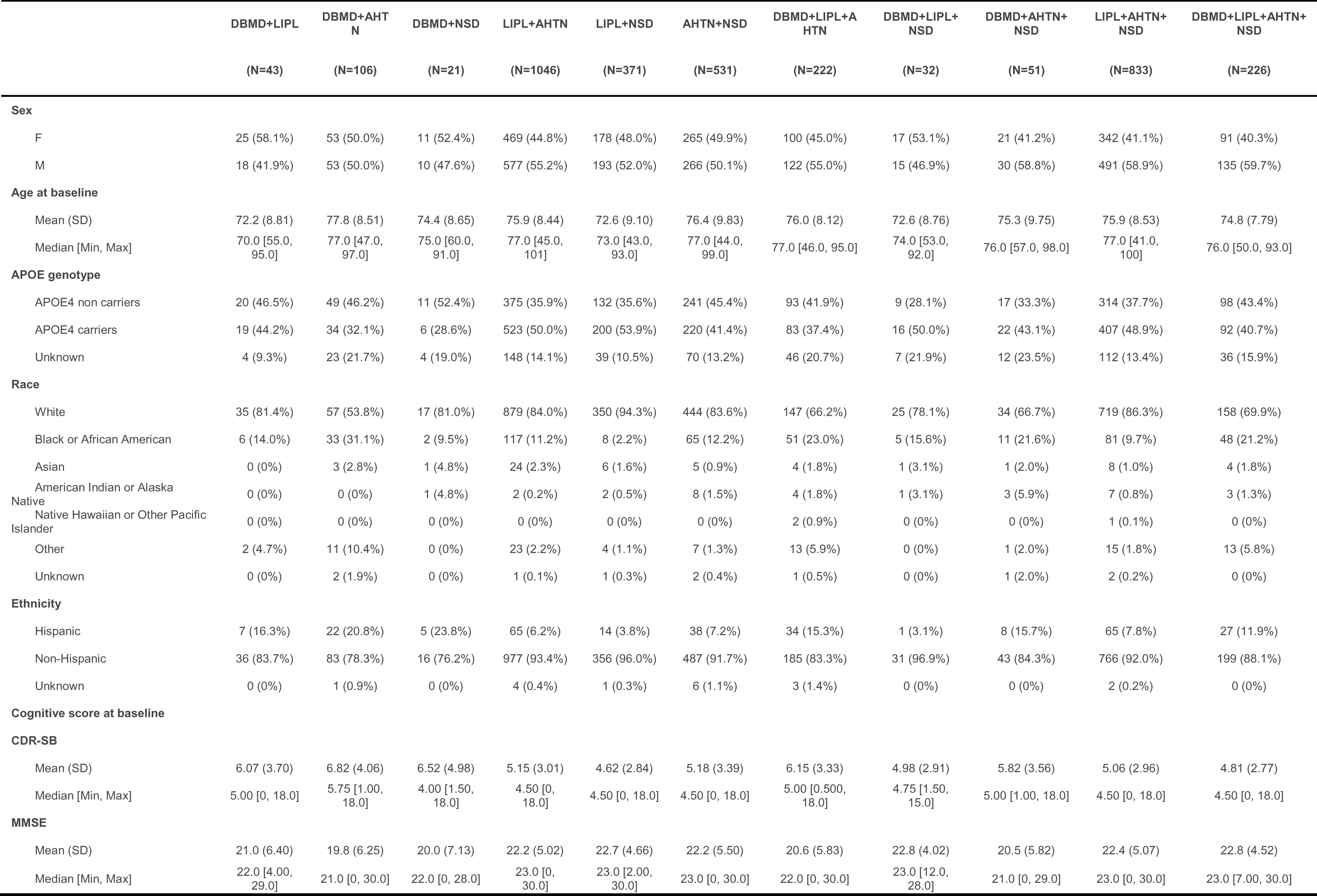
Alzheimer’s Disease NACC participant demographics and baseline cognitive scores by therapeutic group.

### 2.4. Secondary Outcomes

Change in CDR-SB from baseline for each therapeutic group was assessed based on sex and APOE status including APOE4 carriers and non-APOE4 carriers. Drug groups with insufficient participant number in the sex groups (n<50) were excluded from these analysis (DBMD alone, DBMD+AHTN, DBMD+LIPL, DBMD+NSD, **Table 1**).

### 2.5. Statistical Analysis

Statistical analyses were conducted using R (version 4.2.1). Descriptive statistics were generated to characterize the study cohort at their first visits (baseline) when diagnosed with AD utilizing to all study variables. Continuous variables among groups were compared using one-way analysis of variance (ANOVA), while categorical variables were compared using chi-square testing.

To evaluate the impact of each medication and their combinations on cognitive performance, changes in cognition (CDR-SB and MMSE scores) to the participants’ initial visits were calculated. Linear regression models, incorporating random slopes and intercepts, were employed to model the progressions of cognitive scores by time and medication statuses. [40] The changes in cognition scores at 5 years and 10 years were predicted with 95% confidence intervals. The non-treated group was used as a reference, and the percent delay in cognitive decline for other categories was reported. The models were also adjusted for age at recruitment and baseline cognitive scores.

The regression plots were created using the ggplot2 package [41]. The slopes were compared using the emmeans package [40]. To compare the slopes of the regressions, the non-treated group was set as reference. Comparisons of the resulting regression coefficients between the treated and non-treated groups were reported, p-values were adjusted using Dunnett’s test for multiple comparisons.

To evaluate the effects of sex or APOE genotypes on treatment outcomes, linear mixed-effects models with random slopes and intercepts were employed. These models were used to evaluate the improvement in cognitive scores over time, incorporating the variables Sex:medication or APOE:medication status. Pairwise comparisons between slopes were calculated, and the p-values were adjusted using Tukey’s test for multiple comparisons.

## 3. Results

### 3.1. NACC participant profiles

NACC database included 20,946 AD participants, with 7,653 of participants contributing at least two visits and available medication history for the considered therapeutics (**Table 1, overall**). The cohort was balanced for sex (50% females vs 50% males) and was predominately white and non-Hispanic (85% and 92% for race and ethnicity, respectively). APOE genotype was reported for 87% participants, of these 47% were APOE4 carriers. At baseline, the average MMSE and CDR-SB scores were 21.7 and 5.43, respectively, which corresponds to a mild dementia diagnosis (**Table 1**). Of these participants, 5,684 (74%) of participants were prescribed at least one medication including DBMD, LIPL, AHTN, NSD or any combination of these therapies with the 1,969 (26%) had no prescription records for the selected medications. 3,482 participants (45%) were prescribed a combination therapy (**Table 1**). The distribution of participants prescribed to each therapeutic and their combinations is illustrated in Figure 1. The majority of treated participants were either on AHTN alone (19%) or a combination of AHTN and LIPL (18%). The therapeutic groups with the least number of participants (n<50) included the combination of NSD+LIPL+DBMD (0.5%), DBMD alone (0.6%), and DBMD+LIPL (0.7%) (**Figure 1**). Differences in the demographics and baseline cognitive scores between treated and non-treated participants are reported in **Table S3**.

**Figure 1:**
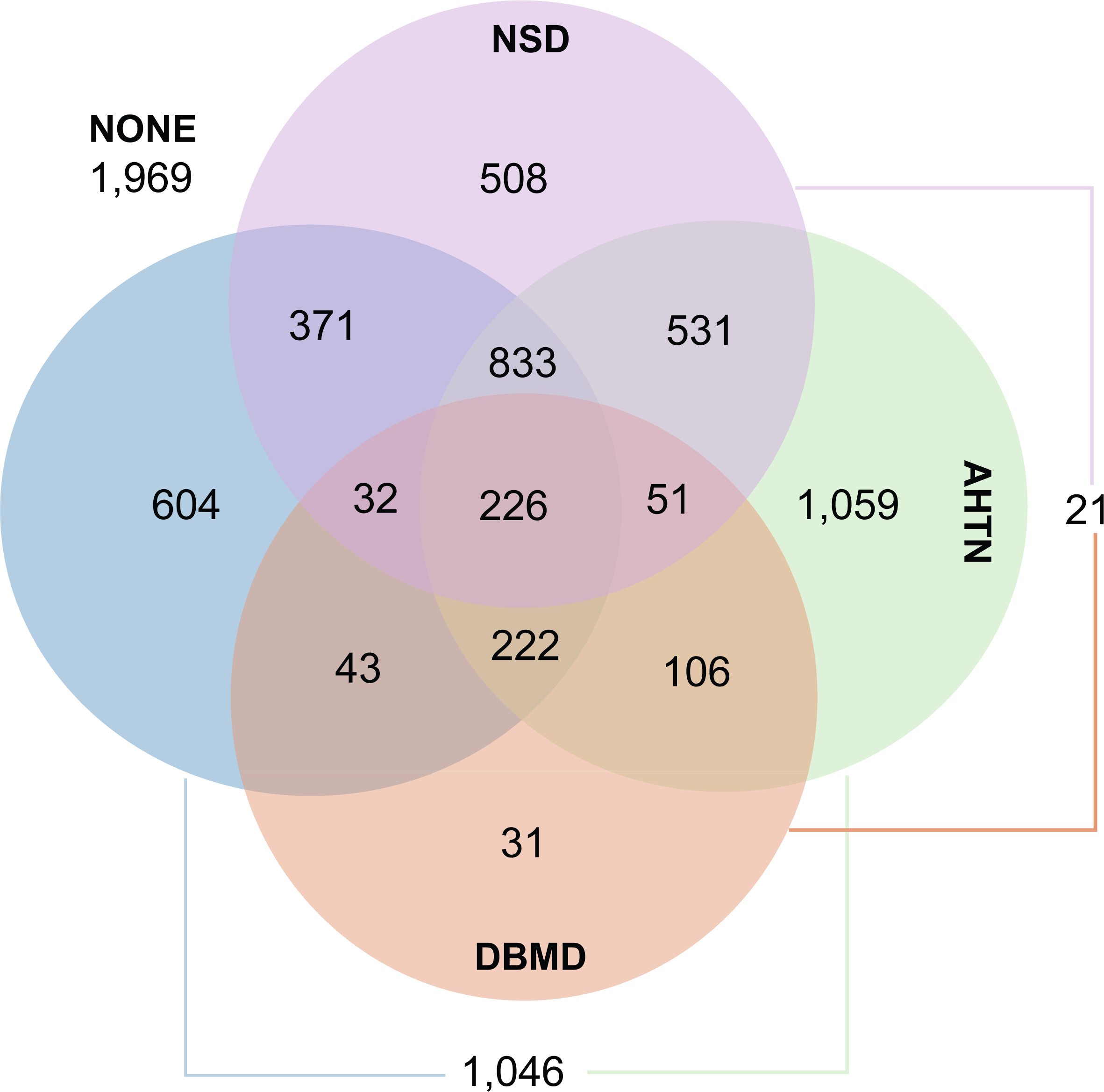
Venn diagram of Alzheimer’s Disease NACC participants distribution across therapeutic groups. The number of participants in each therapeutic group is represented with color overlap (e.g. 226 participants are using DBMD, LIPL, AHTN, and NSD) or lines (e.g. 21 participants are using DBMD and NSD). Abbreviations: DBMD, Diabetes medication; NSD, Non-steroidal anti-inflammatories; LIPL, lipid-lowering drugs; AHTN, antihypertensive drugs.

### 3.2. Impact of therapies prescribed for AD risk factors on cognitive decline

Analyses were conducted evaluating changes in MMSE and CDR-SB scores from baseline over a period of 5 and 10 years for treated and non-treated participants. Results indicated different trajectories of cognitive decline in AD participants at both 5 and 10 years (**Figure 2**, **Table 2**). AD participants not treated with any AD-risk-factor medication exhibited an average decline in MMSE score of –9.2 (95% Confidence Interval [CI], [-9.5, -8.9]) and -17.8 [-18.4, -17.2] points over a period of 5 and 10 years, respectively (**Figure 2a**, **Table 2**), with a 6.9 fold total decline from baseline MMSE levels over 10 years (**Figure 2b**). Ranking of combination therapeutics according to efficacy to delay cognitive decline, resulted in the following ranking in efficacy: (i) DBMD+LIPL+AHTN+NSD, (ii) LIPL+AHTN+NSD, (iii) DBMD+LIPL+AHTN, (iv) DBMD+AHTN+NSD, (v) LIPL+AHTN, (vi) AHTN+NSD, (vii) AHTN, (viii) LIPL+NSD, (ix) LIPL, (x) NSD, (xi) DBMD+AHTN (**Figure 2a**, **Table 2**). The quadruplet combination of DBMD+LIPL+AHTN+NSD exhibited the most favorable outcome, with a 44% and 47% slowing of cognitive decline compared to controls at 5 and 10 years, respectively (MMSE score change: -5.2 [-5.7, -4.6] in 5 years; -9.5 [-10.7, -8.2] in 10 years; p<0.001), and a total decline of 1.7 fold over 10 years (**Figure 2b**).

**Figure 2:**
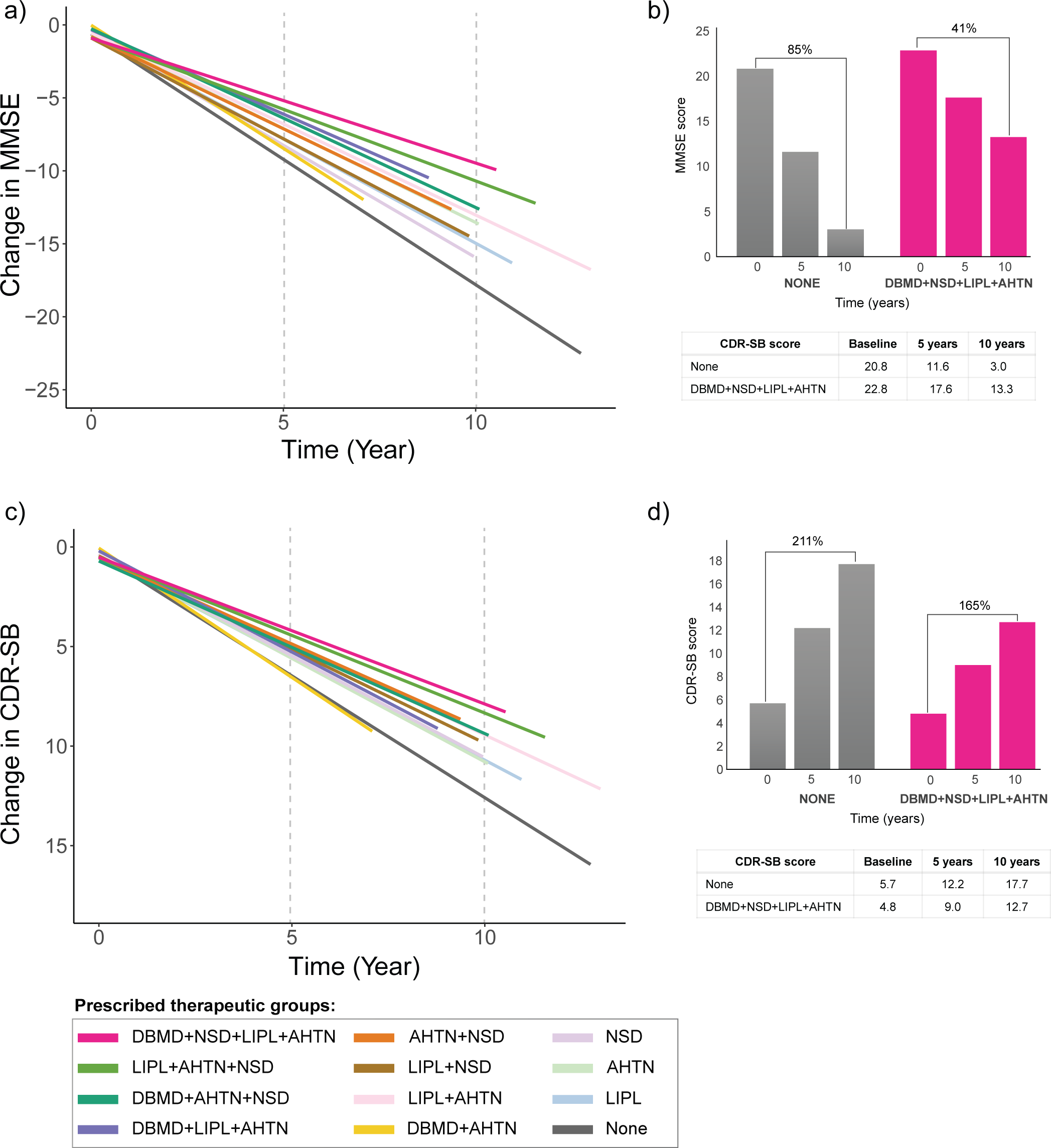
Mini-Mental State Examination (MMSE) and Clinical Dementia Rating Scale Sum of Boxes (CDR-SB) score change from baseline over a 10-year period by therapeutic group. 2a) Change in MMSE score from baseline over 10 years by therapeutic group. 2b) Control (none) and DBMD+NSD+LIPL+AHTN MMSE score at 5 and 10 years and percent change from baseline. 2c) Change in CDR-SB score from baseline over 10 years by therapeutic group. 2d) Control (none) and DBMD+NSD+LIPL+AHTN CDR-SB score at 5 and 10 years and percent change from baseline. Abbreviations: DBMD, Diabetes medication; NSD, Non-steroidal anti-inflammatories; LIPL, lipid-lowering drugs; AHTN, antihypertensive drugs.

**Table 2:**
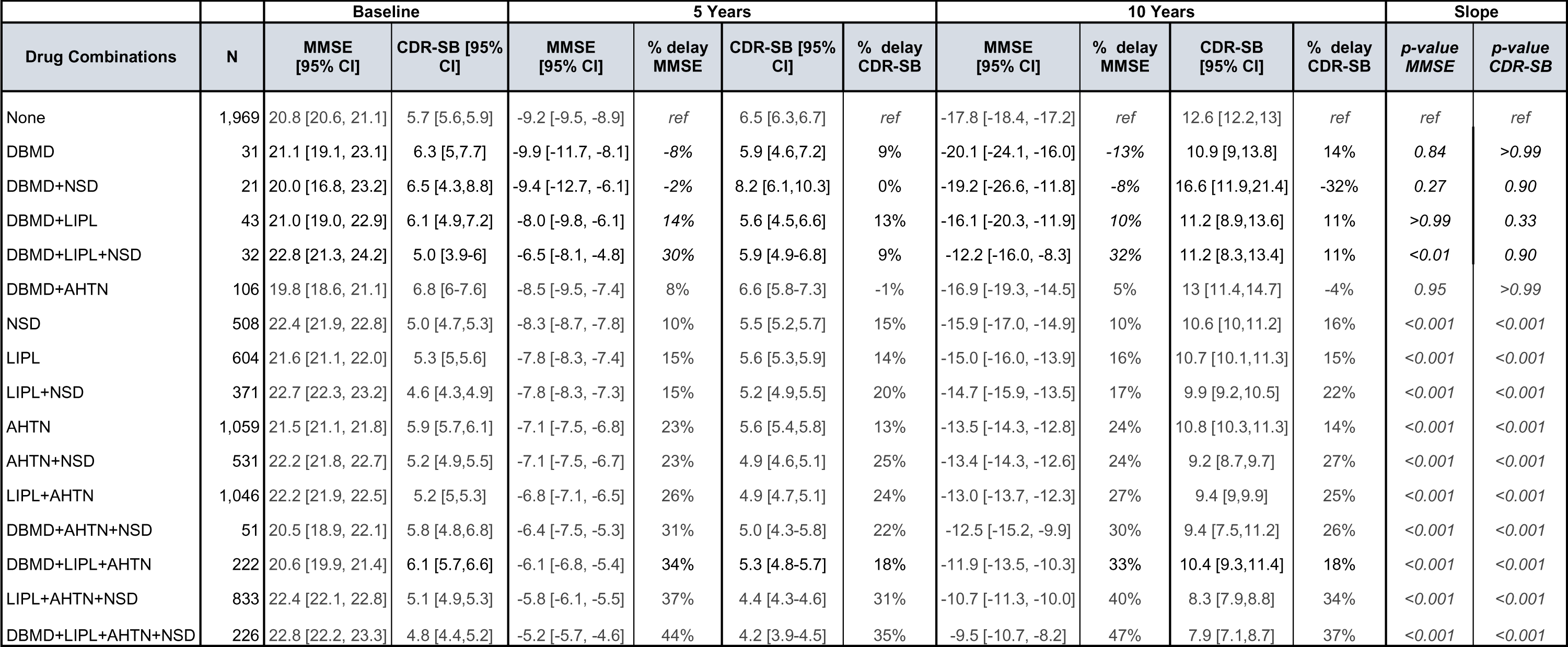
Ranking of drug combination based on Mini-Mental State Examination (MMSE) and Clinical Dementia Rating Scale Sum of Boxes (CDR-SB) score change from baseline for each therapeutic group at baseline, 5 and 10 years.

CDR-SB results were consistent with MMSE outcomes (**Figure 2c**, **Table 2**), in which the quadruplet combination therapy was associated with the greatest delay of cognitive decline compared to control (CDR-SB score change: 4.2 [3.9, 4.5] in 5 years; 7.9 [7.1, 8.7] in 10 years; p<0.001). Ranking of CDR-SB scores was the following: (i) DBMD+LIPL+AHTN+NSD, (ii) LIPL+AHTN+NSD, (iii) AHTN+NSD, (iv) DBMD+AHTN+NSD, (v) LIPL+AHTN, (vi) LIPL+NSD, (vii) DBMD+LIPL+AHT, (viii) NSD, (ix) LIPL, (x) AHTN (**Figure 2c**, **Table 2**). Over the course of 10 years, the untreated group declined 3.1 fold from baseline while participants treated with the quadruplet combination declined 2.6 fold (**Figure 2d**). Results from the adjusted analysis based on age and baseline cognitive scores (MMSE and CDR-SB) yielded comparable results (**Table S4, Table S5**).

### 3.3. Sex differences

Sex differences analysis of cognitive decline associated with each therapeutic prescribed alone or in pairs is shown in **Figure 3**. Non-treated AD participants did not exhibit significant sex differences (p = 0.80) (**Table 3**, **Figure 3**, dashed lines) with both sexes declining at a comparable rate. In contrast, AD participants on AD-risk-factor therapeutics exhibited a sex-dependent magnitude of delay in cognitive decline. Significant sex differences occurred in participants treated with LIPL alone (p<0.001) (**Table 3**, **Figure 3a**), AHTN+NSD (p = 0.03) (**Table 3**, **Figure 3e**), and NSD alone (p<0.01) (**Table 3**, **Figure 3f**), where greater slowing of cognitive decline was associated with male sex. Conversely, AHTN alone did not exhibit a sex-dependent effect on cognition (p>0.99) (**Table 4**, **Figure 3d**); however, when AHTN was combined with NSD, males exhibited slower decline (p=0.03) compared to females (**Table 4**, **Figure 3e**). In contrast, females treated with LIPL alone (**Figure 3a**) did not exhibit a significant difference compared to non-treated participants (p=0.95). However, when in combination LIPL plus, either AHTN or NSD, was associated with significant delay in cognitive decline in females was evident (p<0.001).

**Figure 3:**
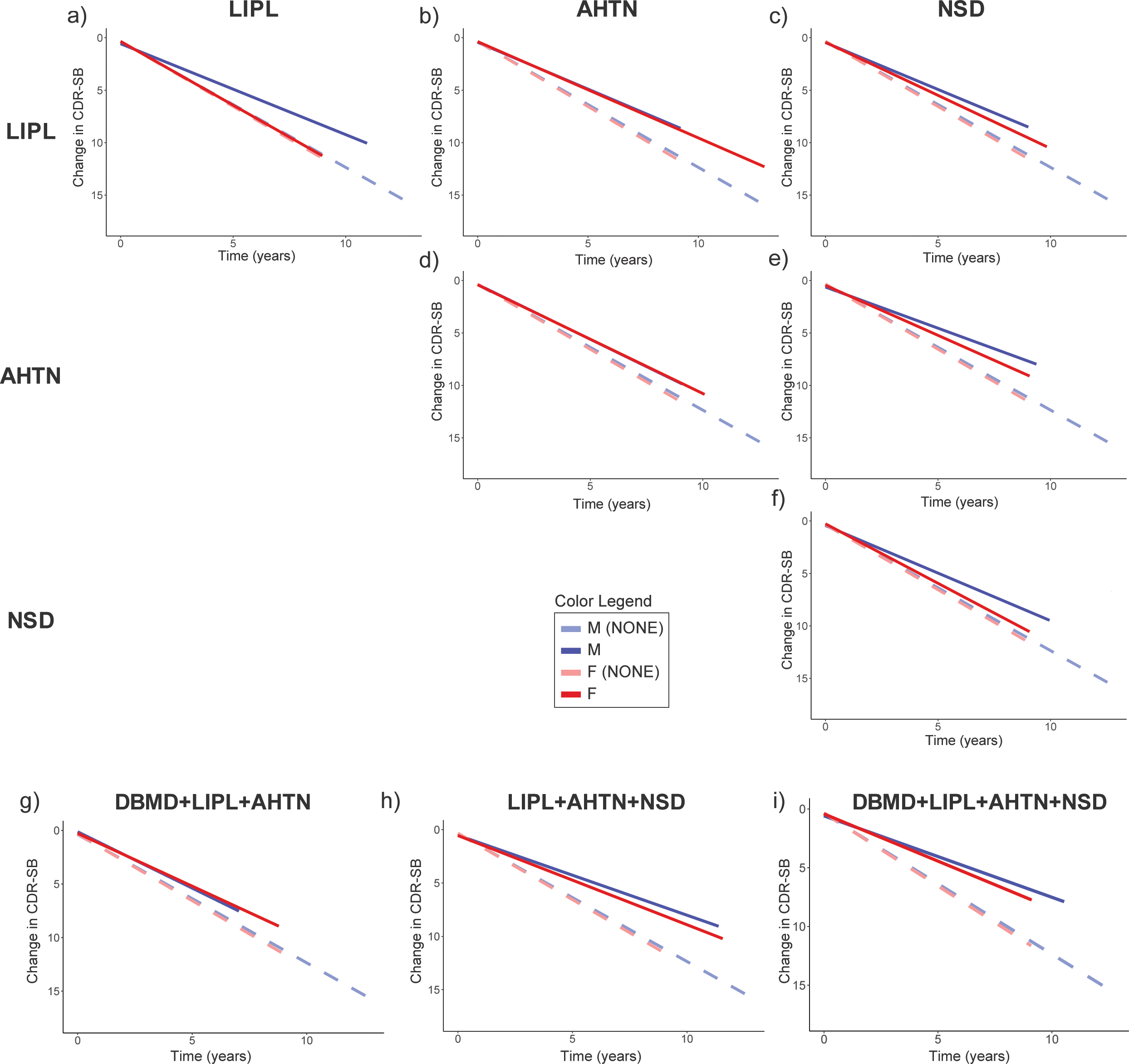
Sex differences in Clinical Dementia Rating Scale Sum of Boxes (CDR-SB) score change from baseline over a 10-year period by drug combination.

**Table 3:**
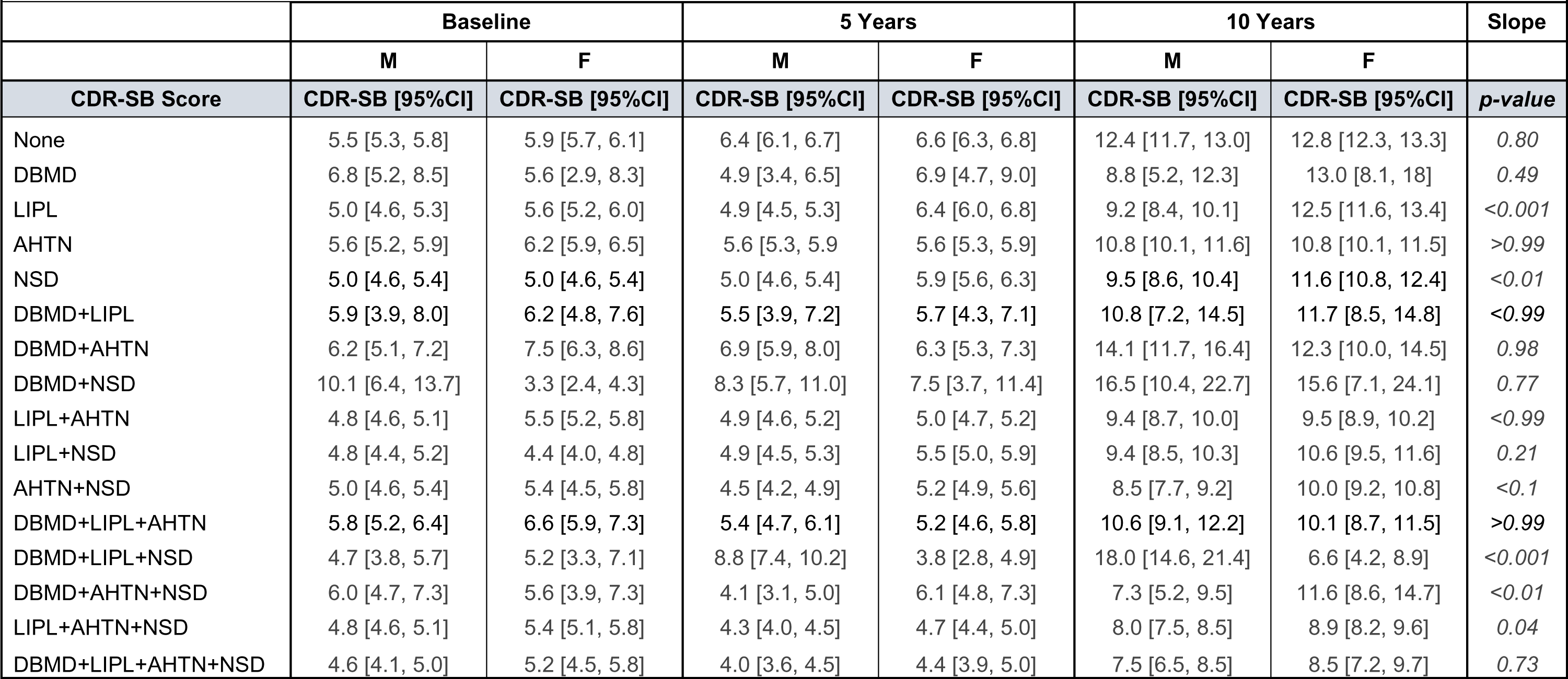
Sex differences in Clinical Dementia Rating Scale Sum of Boxes (CDR-SB) score change from baseline and percent improvement compared to control (none) at 5 and 10 years by drug combination.

| Table 3: Sex differences in CDR-sum of boxes (CDR-SB) score change from baseline compared to control (none) at 5 and 10 years by therapeutic group |  |  |  |  |  |  |  |
| --- | --- | --- | --- | --- | --- | --- | --- |
|  | Baseline |  | 5 Years |  | 10 Years |  | Slope |
|  | M | F | M | F | M | F |  |
| CDR-SB Score | CDR-SB [95%CI] | CDR-SB [95%CI] | CDR-SB [95%CI] | CDR-SB [95%CI] | CDR-SB [95%CI] | CDR-SB [95%CI] | p-value |
| None | 5.5 [5.3, 5.8] | 5.9 [5.7, 6.1] | 6.4 [6.1, 6.7] | 6.6 [6.3, 6.8] | 12.4 [11.7, 13.0] | 12.8 [12.3, 13.3] | 0.80 |
| DBMD | 6.8 [5.2, 8.5] | 5.6 [2.9, 8.3] | 4.9 [3.4, 6.5] | 6.9 [4.7, 9.0] | 8.8 [5.2, 12.3] | 13.0 [8.1, 18] | 0.49 |
| LIPL | 5.0 [4.6, 5.3] | 5.6 [5.2, 6.0] | 4.9 [4.5, 5.3] | 6.4 [6.0, 6.8] | 9.2 [8.4, 10.1] | 12.5 [11.6, 13.4] | <0.001 |
| AHTN | 5.6 [5.2, 5.9] | 6.2 [5.9, 6.5] | 5.6 [5.3, 5.9] | 5.6 [5.3, 5.9] | 10.8 [10.1, 11.6] | 10.8 [10.1, 11.5] | >0.99 |
| NSD | 5.0 [4.6, 5.4] | 5.0 [4.6, 5.4] | 5.0 [4.6, 5.4] | 5.9 [5.6, 6.3] | 9.5 [8.6, 10.4] | 11.6 [10.8, 12.4] | <0.01 |
| DBMD+LIPL | 5.9 [3.9, 8.0] | 6.2 [4.8, 7.6] | 5.5 [3.9, 7.2] | 5.7 [4.3, 7.1] | 10.8 [7.2, 14.5] | 11.7 [8.5, 14.8] | <0.99 |
| DBMD+AHTN | 6.2 [5.1, 7.2] | 7.5 [6.3, 8.6] | 6.9 [5.9, 8.0] | 6.3 [5.3, 7.3] | 14.1 [11.7, 16.4] | 12.3 [10.0, 14.5] | 0.98 |
| DBMD+NSD | 10.1 [6.4, 13.7] | 3.3 [2.4, 4.3] | 8.3 [5.7, 11.0] | 7.5 [3.7, 11.4] | 16.5 [10.4, 22.7] | 15.6 [7.1, 24.1] | 0.77 |
| LIPL+AHTN | 4.8 [4.6, 5.1] | 5.5 [5.2, 5.8] | 4.9 [4.6, 5.2] | 5.0 [4.7, 5.2] | 9.4 [8.7, 10.0] | 9.5 [8.9, 10.2] | <0.99 |
| LIPL+NSD | 4.8 [4.4, 5.2] | 4.4 [4.0, 4.8] | 4.9 [4.5, 5.3] | 5.5 [5.0, 5.9] | 9.4 [8.5, 10.3] | 10.6 [9.5, 11.6] | 0.21 |
| AHTN+NSD | 5.0 [4.6, 5.4] | 5.4 [4.5, 5.8] | 4.5 [4.2, 4.9] | 5.2 [4.9, 5.6] | 8.5 [7.7, 9.2] | 10.0 [9.2, 10.8] | <0.1 |
| DBMD+LIPL+AHTN | 5.8 [5.2, 6.4] | 6.6 [5.9, 7.3] | 5.4 [4.7, 6.1] | 5.2 [4.6, 5.8] | 10.6 [9.1, 12.2] | 10.1 [8.7, 11.5] | >0.99 |
| DBMD+LIPL+NSD | 4.7 [3.8, 5.7] | 5.2 [3.3, 7.1] | 8.8 [7.4, 10.2] | 3.8 [2.8, 4.9] | 18.0 [14.6, 21.4] | 6.6 [4.2, 8.9] | <0.001 |
| DBMD+AHTN+NSD | 6.0 [4.7, 7.3] | 5.6 [3.9, 7.3] | 4.1 [3.1, 5.0] | 6.1 [4.8, 7.3] | 7.3 [5.2, 9.5] | 11.6 [8.6, 14.7] | <0.01 |
| LIPL+AHTN+NSD | 4.8 [4.6, 5.1] | 5.4 [5.1, 5.8] | 4.3 [4.0, 4.5] | 4.7 [4.4, 5.0] | 8.0 [7.5, 8.5] | 8.9 [8.2, 9.6] | 0.04 |
| DBMD+LIPL+AHTN+NSD | 4.6 [4.1, 5.0] | 5.2 [4.5, 5.8] | 4.0 [3.6, 4.5] | 4.4 [3.9, 5.0] | 7.5 [6.5, 8.5] | 8.5 [7.2, 9.7] | 0.73 |

**Table 4:** APOE genotype differences in Clinical Dementia Rating Scale Sum of Boxes (CDR-SB) score change from baseline and percent improvement compared to control (none) at 5 and 10 years by drug combination.

| Table 4: APOE genotype differences in CDR-sum of boxes (CDR-SB) score change from baseline compared to control (none) at 5 and 10 years by therapeutic group |  |  |  |  |  |  |  |
| --- | --- | --- | --- | --- | --- | --- | --- |
|  | Baseline |  | 5 Years |  | 10 Years |  | Slope |
|  | APOE4 non-carriers | APOE4 carriers | APOE4 non-carriers | APOE4 carriers | APOE4 non-carriers | APOE4 carriers |  |
| CDR-SB Score | CDR-SB [95%CI] | CDR-SB [95%CI] | CDR-SB [95%CI] | CDR-SB [95%CI] | CDR-SB [95%CI] | CDR-SB [95%CI] | p-value |
| None | 5.4 [5.2, 5.7] | 5.7 [5.5, 5.9] | 6.1 [5.8, 6.4] | 6.8 [6.6, 7.1] | 11.8 [11.2, 12.5] | 12.1 [11.3, 12.8] | <0.001 |
| DBMD | 5.1 [3.9, 6.4] | 7.2 [3.8, 10.6] | 6.1 [4.2, 8.0] | 5.6 [3.5, 7.6] | 11.0 [6.6, 15.4] | 10.3 [5.6, 15.0] | 0.88 |
| LIPL | 5.5 [5.0, 6.0] | 5.1 [4.8, 5.5] | 4.4 [3.9, 4.8] | 6.5 [6.1, 6.8] | 8.0 [7.1, 9.0] | 12.6 [11.7, 13.5] | <0.001 |
| AHTN | 5.7 [5.4, 6.0] | 5.7 [5.4, 6.1] | 4.7 [4.4, 5.0] | 6.2 [5.9, 6.5] | 8.9 [8.1, 9.6] | 12.1 [11.3, 12.8] | <0.001 |
| NSD | 4.6 [4.2, 5.0] | 5.0 [4.6, 5.4] | 5.3 [4.9, 5.7] | 5.4 [5.1, 5.8] | 10.4 [9.4, 11.3] | 10.4 [9.6, 11.3] | 0.86 |
| DBMD+LIPL | 5.5 [3.7, 7.3] | 5.9 [4.7, 7.1] | 6.9 [4.8, 8.9] | 5.1 [3.9, 6.3] | 14.0 [9.4, 18.6] | 10.1 [7.4, 12.8] | 0.70 |
| DBMD+AHTN | 6.4 [5.3, 7.5] | 7.0 [5.5, 8.5] | 8.1 [6.9, 9.3] | 6.0 [5.1, 7.0] | 16.1 [13.4, 18.9] | 11.9 [9.7, 14.1] | 0.04 |
| DBMD+NSD | 5.2 [3.1, 7.3] | 6.3 [-0.5, 13.2] | 9.2 [6.4, 12.0] | 4.3 [-1.0,9.5] | 18.7 [12.5, 25] | 8.9 [-2.9, 20.8] | 0.10 |
| LIPL+AHTN | 4.9 [4.6, 5.2] | 5.1 [4.9, 5.4] | 4.4 [4.1, 4.8] | 5.1 [4.8, 5.4] | 8.4 [7.6, 9.2] | 9.9 [9.3, 10.5] | 0.04 |
| LIPL+NSD | 4.4 [3.9, 4.9] | 4.6 [4.2, 5.0] | 4.4 [3.9, 4.9] | 5.6 [5.3, 6.0] | 8.0 [6.9, 9.2] | 11.0 [10.1, 11.9] | <0.01 |
| AHTN+NSD | 5.1 [4.7, 5.5] | 5.7 [5.5, 5.9] | 4.4 [4.0, 4.7] | 5.1 [4.8, 5.4] | 8.0 [7.2, 8.8] | 9.7 [9.0, 10.4] | 0.04 |
| DBMD+LIPL+AHTN | 6.2 [5.5, 6.9] | 5.4 [4.8, 6.1] | 5.0 [4.3, 5.8] | 5.3 [4.6, 6.0] | 10.0 [8.3, 11.6] | 10.4 [8.8, 12.0] | 0.87 |
| DBMD+LIPL+NSD | 4.2 [2.3, 6.1] | 4.5 [3.6, 5.4] | 4.8 [3.4, 6.1] | 7.0 [5.5, 8.5] | 8.6 [5.3, 12.0] | 13.5 [10.1, 17.0] | 0.11 |
| DBMD+AHTN+NSD | 5.5 [4.1, 6.9] | 5.7 [4.0, 7.4] | 4.6 [3.5, 5.6] | 6.1 [4.9, 7.4] | 8.2 [5.6, 10.7] | 8.0 [8.8, 14.8] | 0.15 |
| LIPL+AHTN+NSD | 4.8 [4.5, 5.1] | 5.0 [4.7, 5.2] | 4.4 [4.1, 4.7] | 4.3 [4.1, 4.6] | 8.3 [7.6, 9.0] | 8.1 [7.5, 8.6] | >0.99 |
| DBMD+LIPL+AHTN+NSD | 4.6 [4.1, 5.2] | 5.0 [4.4, 5.6] | 3.7 [3.2, 4.2] | 4.5 [4.0, 5.1] | 6.7 [5.6, 7.9] | 8.6 [7.4, 9.9] | 0.09 |

Results of sex differences analysis evaluating the impact of combinations of three or four drugs on CDR-SB change are represented in **Figure 3g-i**. The triple combination of LIPL+AHTN+NSD (**Figure 3h**) exhibited significant delay of cognitive decline in both sexes with males exhibiting greater improvement relative to females (p=0.04) (**Table 3**). The quadruplet combination of DBMD+LIPL+AHTN+NSD (**Figure 3i**) exhibited significant delay in cognitive decline in both females and males (p<0.001) and was equally effective for both sexes (p=0.73) (**Table 3**).

### 3.4. APOE genotype differences

Secondary analysis evaluating cognitive decline associated with each therapeutic group by APOE genotype is presented in **Figure 4a-i**. In the non-treated population, APOE4 carriers exhibited greater rate of cognitive decline compared to APOE4 non-carriers (p<0.001) (**Table 4**, **Figure 4**, dashed lines). Participants treated with LIPL alone (**Figure 4a**), AHTN alone (**Figure 4d**), LIPL+AHTN (**Figure 4b**), LIPL+NSD (**Figure 4c**), and HTN+NSD (**Figure 4e**) exhibited significant APOE genotype differences (p<0.05), where APOE4 non-carriers exhibited slower cognitive decline compared to APOE4 carriers (**Table 4**). Furthermore, APOE4 carriers treated with LIPL alone (**Figure 4a**) did not exhibit substantial differences from non-treated participants (p=0.39). However, when these drugs were combined with NSD or AHTN significant delay of cognitive decline in APOE4 carriers was evident (p<0.001).

**Figure 4:**
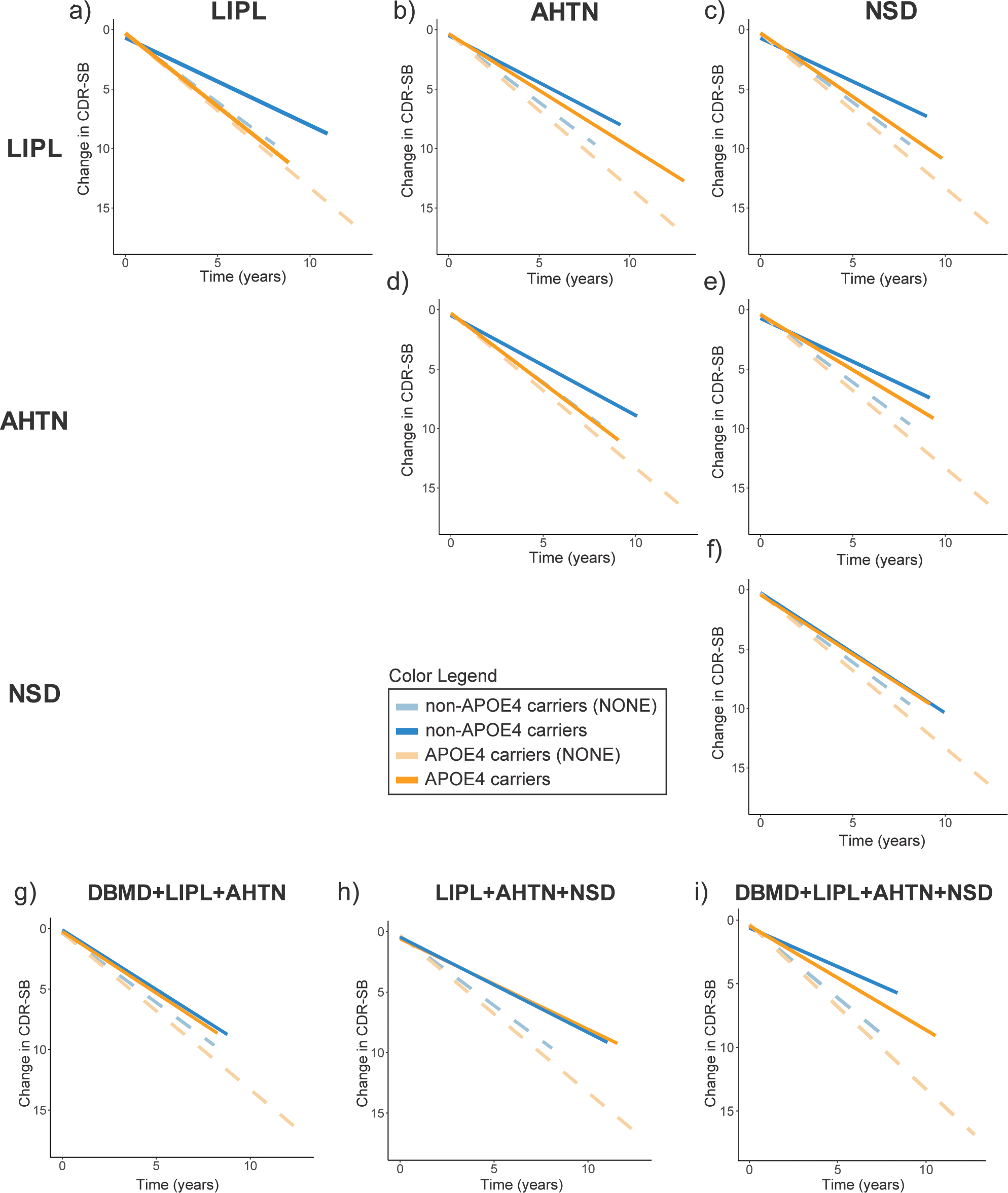
APOE genotype differences in Clinical Dementia Rating Scale Sum of Boxes (CDR-SB) score change from baseline over a 10-year period by drug combination.

## 4. Discussion

The impact of DBMD, LIPL, AHTN, and NSD on risk of developing AD is well documented [13-20], as is their failure to effectively treat AD [21-34]. Single-target randomized clinical trials of risk-factor therapeutics for non-symptomatic AD patients were not successful [21-34]. The disparity between outcomes of epidemiological and medical informatic analyses reporting reduced risk of AD vs the consistent failure to effectively treat or cure AD with these same preventative drug classes has been attributed to the lack of predictive validity of real-world medical treatment data. Herein, we addressed the hypothesis that these therapeutics, when prescribed to AD individuals for their intended purpose, may exert therapeutic benefit by conducting a longitudinal analysis to assess efficacy of DBMD, LIPL, AHTN, and NSD as treatment under real-world conditions in patients diagnosed with AD and prescribed therapeutic interventions for their intended purpose.

Analyses of therapeutic profiles of NACC participants revealed that 74% of AD individuals were treated with at least one therapeutic, underlining the prevalence of risk factors in individuals diagnosed with AD. Furthermore, 61% of treated participants received a combination of drugs rather which is consistent with coexistence of AD risk factors and the multiple systems of biology associated with risk of AD. For example, only 0.6% of the treated population was exclusively prescribed DBMD, while 18% were prescribed a combination including DBMD. These data highlight the complexity and heterogeneity of different therapeutic approaches for addressing the dysregulated systems of biology implicated in AD.

The cognitive performance of AD participants treated with monotherapy of LIPL, AHTN, or NSD as assessed by MMSE and CDR-SB was significantly delayed (10% to 24%) compared to untreated participants at 10 years. Furthermore, the data indicated additive and synergistic efficacy in slowing rate of cognitive decline. The most significant delaying of decline, reaching 47% and 37% at 10 years considering MMSE and CDR-SB scores, respectively, was observed in individuals prescribed with the combination of DBMD+LIPL+AHTN+NSD, while other combinations of two or three drugs reached a delay of cognitive decline ranging from 15% to 40% compared to control. This finding supports a combination approach targeting multiple AD risk factors which could be more effective than monotherapy, given the multiple systems of biology, including risk factors, disrupted in AD patients.

Notably, individuals on combination therapies had better cognitive performance at baseline and were older at time of enrollment. These findings are consistent with combination therapy sustaining cognitive function in those at risk for AD and delaying age of onset.

Secondary analyses conducted to evaluate the influence of sex and APOE genotype on cognitive decline in treated AD participants revealed that sex and APOE genotype impacted magnitude of decline. This finding supports clinical studies that identified a subset of responders within the unsuccessful clinical trials of these therapies, emphasizing the role of sex and APOE genotype in treatment outcomes [35-37]. In cases where a specific sex or genotype did not achieve cognitive improvement through a singular therapeutic intervention, the introduction of a supplementary second or third drug led to a significant delay in cognitive decline; highlighting the therapeutic potential of combination therapy for non-responder individuals to single-treatment approaches. Moreover, these data emphasize the need for integrating sex and APOE genotype as precision medicine strategies in the development of effective treatments for AD.

A strength of the NACC data is the real-world AD clinical heterogeneity which contrasts with the more harmonized participant groups of randomized controlled clinical trials that are selected with stringent inclusion and exclusion criteria. A limitation of the NACC data is the number of participants who met the criteria for inclusion in this study, 7,653 AD participants, with some therapeutic groups including fewer than 50 participants. Another limitation of NACC data is the self-reported nature of medication use data of participants which was not confirmed by a clinician. Outcomes of our analysis provide a therapeutic class framework from which identification of specific drugs with greatest efficacy within each class should be determined for optimized precision combination therapies using larger and comprehensive datasets.

Overall, these findings indicate the potential of FDA-approved drugs targeting AD risk factors to significantly modify rate of cognitive decline. It is noteworthy that the magnitude of delay in cognitive decline of effective therapeutic combinations reported herein was comparable to, if not greater than recently approved amyloid antibodies lecanemab and aducanumab. Specifically, the quadruplet combination of DBMD+LIPL+AHTN+NSD was associated with a 35% and 37% (CDR-SB adjusted mean difference vs. placebo: -2.30 at 5 years and -4.7 at 10 years) slowing of CDR-SB cognitive decline at 5 and 10 years, respectively, whereas lecanemab was associated with 27% slowing of CDR-SB cognitive decline over 18 months (CDR-SB adjusted mean difference vs. placebo: -0.45) [7] and aducanumab achieved 22% slowing of cognitive decline in 78 weeks (CDR-SB adjusted mean difference vs. placebo: -0.39) [8]. Notably, patients within the lecanemab ([mean age]: 71 years old) and aducanumab [70 years old] clinical trials had baseline CDR-SB scores of approximately 3.2 and 2.4, respectively [7, 8], which represent a stage of “questionable impairment” (0.5-4.0) [42]. Whereas NACC participants were older [74 years old] and had a mean CDR-SB baseline score of 5.4 indicative of a stage of mild dementia (4.5-9.0) [42]. As neither of these approaches effectively reverse the progression of the disease, it is critical to identify mechanisms not targeted by these therapeutic approaches like brain atrophy and/or neurofibrillary tangles. Nevertheless, significantly delaying rate of cognitive decline has clinical benefit for AD patients and could extend a window of therapeutic opportunity for reversal of disease.

In conclusion, combination therapies targeting AD risk factors significantly slowed cognitive decline in AD participants at a magnitude comparable to or greater than beta-amyloid immunomodulator interventions. These data support combination precision medicine targeting AD risk factors to significantly delay the course of the disease that persists for a decade.

## Data Availability

All data produced in the present study are available upon reasonable request to the authors

## Acknowledgments/Funding Sources

Research reported herein was supported by the National Institute on Aging (grants P01AG026572 [Perimenopause in Brain Aging and Alzheimer’s Disease], 5R01AG057931-02 [Sex Differences in Molecular Dementias of Alzheimer’s Disease Risk: Prodromal Endophenotype]), the Women’s Alzheimer’s Movement to Roberta Diaz Brinton, and the University of Arizona Center for Innovation in Brain Science.

## Authors’ contributions

Yuan Shang: Formal analysis (lead); Methodology (lead); Writing-original draft (lead). Georgina Torrandell-Haro: Formal analysis (lead); Methodology (supporting); Writing-original draft (lead). Francesca Vitali: Methodology (supporting); Writing-original draft (supporting); Writing-review & editing (equal); Supervision (equal). Roberta Diaz Brinton: Conceptualization (lead); Writing-review & editing (equal); Funding acquisition (lead); Supervision (lead).

## Consent Statement

This study was conducted using de-identified data provided by the National Alzheimer’s Coordinating Center (NACC). No additional data were collected from participants without their informed consent.

## Availability of data and materials

The database used in this study are available from: National Alzheimer’s Coordinating Center (NACC). These data are available upon request.

