## Supplementary material for "Combination therapies delay cognitive decline over 10 years in Alzheimer’s NACC participants": Table S1

### Supplemental Materials

| *Table S1: Codes used* | | |
|---|---|---|
| **Form** | **Variable** | **Code name** |
| Subject Demographics (A1) | Year of birth | BIRTHYR |
| | Sex | SEX |
| | Hispanic/Latino ethnicity | HISPANIC |
| | Race | RACE |
| Subject Medications (A4) | Reported current use of lipid lowering medication | NACCLIPL |
| | Reported current use of any type of antihypertensive or blood pressure medication | NACCHTN |
| | Reported current use of nonsteroidal anti-inflammatory medication | NACCNSD |
| | Reported current use of a diabetes medication | NACCDBMD |
| Clinician Diagnosis | Alzheimer's Disease diagnosis | NACCUDSD |
| Neuropsychological Battery Summary Scores (C1) | Total MMSE score | NACCMMSE |
| CDR Plus NACC FTDL (B4) | CDR sum of boxes | CDRSUM |
