## Supplementary material for "Combination therapies delay cognitive decline over 10 years in Alzheimer’s NACC participants": Table S2

| ***Table S2:*** *Medication codes used and drugs included in each category* | | |
|---|---|---|
| **Reported Medication** | **Code** | **Drugs** |
| Lipid lowering medication (LIPL) | NACCLIPL | lovastatin, pravastatin, simvastatin, fluvastatin, atorvastatin, cerivastatin, red yeast rice, rosuvastatin, pitavastatin, niacin, probucol, dextrothyroxine sodium, icosapent ethyl, clofibrate, gemfibrozil, fenofibrate, fenofibric acid, cholestyramine, colestipol, colesevelam, ezetimibe, lovastatin-niacin, aspirin-pravastatin, amlodipine-atorvastatin, ezetimibe-simvastatin, niacin-simvastatin, simvastatin-sitagliptin |
| Nonsteroidal anti-inflammatory medication (NSD) | NACCNSD | ibuprofen, naproxen, fenoprofen, ketoprofen, sulindac, indomethacin, tolmetin, flurbiprofen, ketorolac, meclofenamate, mefenamic acid, nabumetone, phenylbutazone, piroxicam, diclofenac, etodolac, oxaprozin, bromfenac, diclofenac-misprostol, meloxicam, lansoprazole-naproxen, esomeprazole-naproxen, famotidine-ibuprofen, aspirin, diflunisal, choline salicylate, salsalate, sodium salicylate, sodium thiosalicylate, magnesium salicylate, choline salicylate-magnesium salicylate, ASA/citric acid/Na bicarb, Al hydroxide/ASA/Ca carbonate/Mg hydoxide, celecoxib, rofecoxib, valdecoxib, APAP/ASA/caffeine/salicylamide, APAP/ASA/Caffeine, APAP/Al hydroxide/ASA/caffeine/Mg hydroxide, ASA/caffeine/salicylamide, aspirin-meprobamate, aspirin-caffeine, aspirin-phenyltoloxamine, magnesium salicylate-phenyltoloxamine, ASA/butalbital/caffeine, aspirin-butalbital, aspirin-diphenydramine, diphenydramine-magnesium salicylate, acetaminophen-salicylamide, APAP/caffeine/phenyltoloxamine/salicylamide, APAP/phenyltoloxamine/salicylamide, APAP/caffeine/mg salicylate/phenyltoloxamin, diphenhydramine-ibuprofen, APAP/caffeine/magnesium salicylate, acetaminophen-aspirin, caffeine-magnesium salicylate, APAP/magnesium salicylate/pamabrom |
| Diabetes medication (DBMD) | NACCDBMD | chlorpropamide, acetohexamide, glipizide, glyburide, tolazamide, tolbutamide, glimepiride, metformin, insulin, other forms of insulin, acarbose, miglitol, troglitazone, rosiglitazone, pioglitazone, repaglinide, nateglinide, glyburide-metformin, metformin-rosiglitazone, glipizide-metformin, metformin-pioglitazone, glimepiride-rosiglitazone, glimepiride-pioglitazone, metformin-sitagliptin, metformin-repaglinide, metformin-saxagliptin, simvastatin-sitagliptin, linagliptin-metformin, sitagliptin, saxagliptin, linagliptin, pramlintide, exenatide, liraglutide |
| Antihypertensive medication (AHTN) | NACCHTN | captopril, enalapril, fosinopril, quinapril, ramipril, benazepril, lisinopril, moexipril, trandolapril, perindopril, atenolol, acebutolol, metoprolol, betaxolol, esmolol, bisoprolol, nebivolol, labetalol, nadolol, propranolol, pindolol, timolol, penbutolol, sotalol, carteolol, carvedilol, diltiazem, verapamil, nifedipine, felodipine, isradipine, nicardipine, nimodipine, bepridil, amlodipine, mibefradil, clevidipine, nisolpidine, furosemide, bumetanide, ethacrynic acid, torsemide, amiloride, spironolactone, triamterene, chlorothiazide, hydrochlorotiazide, indapamide, metolazone, bendroflumethiazide, methylchlorhiazide, benzthiazide, hydroflumethiazide, polythiazide, acetazolamide, dichlorphenamide, mannitol, pamabrom, urea, hydralazine, minoxidil, nitroprusside, nitroglycerin, alprostadil, nesiritide, losartan, valsartan, irbesartan, eprosartan, candesartan, telmisartan, olmesartan, azilsartan, guanethidine, prazosin, reserpine, terazosin, guanadrel, doxazosin, mecamylamine, rauwolfia serpentina, deserpidine, tamsulosin, alfusozin, silodosin, dutasteride-tamsulosin, clonidine, guanabenz, methyldopa, guanfacine. |
