## Supplementary material for "Combination therapies delay cognitive decline over 10 years in Alzheimer’s NACC participants": Table S3

***Table S3:*** *p-values of NACC participant demographics compared to control group (none)*

| | DBMD | LIPL | AHTN | NSD | DBMD+LIPL | DBMD+ AHTN | DBMD+ NSD | LIPL+ AHTN | LIPL+ NSD | AHTN +NSD | DBMD+ LIPL+ AHTN | DBMD +LIPL+ NSD | DBMD+ AHTN+ NSD | LIPL+ AHTN+ NSD | DBMD+LIPL+ AHTN+NSD |
|---|---|---|---|---|---|---|---|---|---|---|---|---|---|---|---|
| **Sex** | 0.195 | 0.390 | 0.895 | 0.979 | 0.920 | 0.602 | 0.972 | **<0.001** | 0.045 | 0.112 | 0.018 | 0.978 | 0.147 | **<0.001** | **<0.001** |
| **Age** | 0.019 | 0.537 | **<0.001** | 0.384 | 0.881 | **<0.001** | 0.477 | **<0.001** | 0.133 | **<0.001** | **<0.001** | 0.826 | 0.054 | **<0.001** | **<0.001** |
| **APOE genotype** | 0.234 | 0.079 | 0.369 | 0.272 | 0.891 | 0.034 | 0.600 | 0.357 | 0.132 | 0.209 | **0.026** | 0.608 | 0.383 | 0.876 | 0.554 |
| **Race** | 0.065 | 0.535 | **<0.001** | 0.339 | 0.952 | **<0.001** | 0.879 | 0.138 | 0.113 | 0.183 | **<0.001** | 0.890 | **0.001** | 0.881 | **<0.001** |
| **Ethnicity** | 0.895 | 0.475 | 0.991 | 0.200 | 0.416 | **<0.001** | 0.143 | 0.447 | 0.064 | 0.561 | **0.003** | 0.873 | 0.404 | 0.902 | 0.289 |
| **CDR-SB** | 0.653 | 0.026 | 0.541 | **<0.001** | 0.837 | **0.012** | 0.611 | **<0.001** | **<0.001** | **0.005** | 0.269 | 0.494 | 0.987 | **<0.001** | **<0.001** |
| **MMSE** | 0.979 | 0.035 | 0.024 | **<0.001** | 0.992 | 0.287 | 0.831 | **<0.001** | **<0.001** | **<0.001** | 0.899 | 0.213 | 0.935 | **<0.001** | **<0.001** |
