## Supplementary material for "Combination therapies delay cognitive decline over 10 years in Alzheimer’s NACC participants": Table S4

*Table S4: Adjusted* ranking of drug combination based on MMSE score change from baseline for each therapeutic group at baseline, 5 and 10 years*

| | | Baseline | 5 Years | | 10 Years | | Slope |
|---|---|---|---|---|---|---|---|
| **Drug Combinations** | **N** | **MMSE (95% CI)** | **MMSE (95% CI)** | **% improvement MMSE** | **MMSE (95% CI)** | **% improvement MMSE** | ***p-value MMSE*** |
| None | 1,969 | 20.8 [20.6, 21.1] | -9.1 [-9.4, -8.9] | *ref* | -17.6 [-18.2, -17.0] | *ref* | *ref* |
| DBMD+AHTN | 106 | 19.8 [18.6, 21.1] | -8.0 [-9.1, -6.8] | 13% | -15.3 [-17.6, -12.9] | 13% | *0.73* |
| NSD | 508 | 22.4 [21.9, 22.8] | -8.3 [-8.7, -7.9] | 10% | -15.9 [-16.6, -15.1] | 10% | *0.01* |
| LIPL+NSD | 371 | 22.7 [22.3, 23.2] | -8.0 [-8.4, -7.5] | 13% | -15.3 [-16.2, -14.4] | 13% | *0.02* |
| LIPL | 604 | 21.6 [21.1, 22.0] | -7.8 [-8.3, -7.4] | 14% | -15.0 [-15.9, -14.0] | 15% | *<0.001* |
| AHTN+NSD | 531 | 22.2 [21.8, 22.7] | -7.2 [-7.5, -6.8] | 21% | -13.7 [-14.4, -13.0] | 22% | *<0.001* |
| AHTN | 1,059 | 21.5 [21.1, 21.8] | -7.1 [-7.4, -6.7] | 23% | -13.4 [-14.1, -12.6] | 24% | *<0.001* |
| LIPL+AHTN | 1,046 | 22.2 [21.9, 22.5] | -6.7 [-7.0, -6.3] | 27% | -12.7 [-13.4, -12.0] | 28% | *<0.001* |
| DBMD+AHTN+NSD | 51 | 20.5 [18.9, 22.1] | -6.2 [-7.2, -5.3] | 32% | -11.8 [-13.8, -9.9] | 33% | *<0.001* |
| DBMD+LIPL+AHTN | 222 | 20.6 [19.9, 21.4] | -5.8 [-6.5, -5.0] | 37% | -10.9 [-12.3, -9.4] | 38% | *<0.001* |
| LIPL+AHTN+NSD | 833 | 22.4 [22.1, 22.8] | -5.8 [-6.1, -5.5] | 37% | -10.9 [-11.5, -10.3] | 38% | *<0.001* |
| DBMD+LIPL+AHTN+NSD | 226 | 22.8 [22.2, 23.3] | -5.1 [-5.6, -4.6] | 45% | -9.5 [-10.5, -8.4] | 46% | *<0.001* |

**adjusted based on age and baseline cognitive scores*
