## Supplementary material for "Combination therapies delay cognitive decline over 10 years in Alzheimer’s NACC participants": Table S5

***Table S5:*** *Adjusted* ranking of drug combination based on CDR-SB score change from baseline for each therapeutic group at baseline, 5 and 10 years*

| | | Baseline | 5 Years | | 10 Years | | Slope |
|---|---|---|---|---|---|---|---|
| **Drug Combinations** | **N** | **CDR-SB (95% CI)** | **CDR-SB (95% CI)** | **% improvement CDR-SB** | **CDR-SB (95% CI)** | **% improvement CDR-SB** | ***p-value CDR-SB*** |
| None | 1,969 | 5.7 [5.6, 5.9] | 6.3 [6.1, 6.5] | *ref* | 12.1 [11.8, 12.5] | *ref* | *ref* |
| DBMD+AHTN | 106 | 6.8 [6.0, 7.6] | 6.2 [5.5, 6.9] | 2% | 11.9 [10.5, 13.4] | 2% | *>0.99* |
| AHTN | 1,059 | 5.9 [5.7, 6.1] | 5.5 [5.2, 5.7] | 13% | 10.5 [10.0, 10.9] | 14% | *<0.001* |
| LIPL | 604 | 5.3 [5, 5.6] | 5.4 [5.2, 5.7] | 13% | 10.5 [9.9, 11.0] | 14% | *<0.001* |
| NSD | 508 | 5.0 [4.7, 5.3] | 5.3 [5.1, 5.5] | 16% | 10.2 [9.7, 10.7] | 16% | *<0.001* |
| LIPL+NSD | 371 | 4.6 [4.3, 4.9] | 5.1 [4.8, 5.4] | 19% | 9.8 [9.2, 10.3] | 19% | *<0.001* |
| DBMD+AHTN+NSD | 51 | 5.8 [4.8, 6.8] | 5.1 [4.5, 5.6] | 20% | 9.7 [8.5, 10.9] | 20% | *<0.001* |
| DBMD+LIPL+AHTN | 222 | 6.1 [5.7, 6.6] | 5.0 [4.6, 5.5] | 20% | 9.6 [8.7, 10.5] | 21% | *<0.001* |
| AHTN+NSD | 531 | 5.2 [4.9, 5.5] | 4.8 [4.6, 5.0] | 23% | 9.2 [8.8, 9.7] | 24% | *<0.001* |
| LIPL+AHTN | 1,046 | 5.2 [5.0, 5.3] | 4.8 [4.6, 5.0] | 24% | 9.2 [8.7, 9.6] | 24% | *<0.001* |
| LIPL+AHTN+NSD | 833 | 5.1 [4.9, 5.3] | 4.3 [4.1, 4.5] | 31% | 8.2 [7.8, 8.5] | 32% | *<0.001* |
| DBMD+LIPL+AHTN+NSD | 226 | 4.8 [4.4, 5.2] | 4.3 [4.0, 4.6] | 32% | 8.1 [7.5, 8.8] | 33% | *<0.001* |

**adjusted based on age and baseline cognitive scores*
